# Human Antibodies Targeting a Transporter Mediate Protection Against Tuberculosis

**DOI:** 10.1101/2020.05.17.20101451

**Authors:** Avia Watson, Hao Li, Bingting Ma, Ronen Weiss, Danielle Bendayan, Lilach Abramovitz, Michael Mor, Erica Pinko, Michal Bar-Oz, Zhenqi Wang, Fengjiao Du, Yu Lu, Jan Rybniker, Hairong Huang, Daniel Barkan, Ye Xiang, Babak Javid, Natalia T Freund

**Affiliations:** Department of Clinical Microbiology and Immunology, Sackler Faculty of Medicine, Tel Aviv University, 6997801 Israel; Centre for Global Health and Infectious Diseases, Collaborative Innovation Centre for the Diagnosis and Treatment of Infectious Diseases, Tsinghua University School of Medicine, Beijing 100084, China; Advanced Innovation Center for Structural Biology, Tsinghua University School of Medicine, Beijing 100084, China; Pulmonary and Tuberculosis Department, Shmuel Harofe Hospital, 70300, Israel; Koret School of Veterinary Medicine, The Robert H. Smith Faculty of Agriculture, Food and Environment, The Hebrew University of Jerusalem, Rehovot, 7610001, Israel; College of Veterinary Medicine, China Agricultural University, Beijing 100094, China; National Clinical Laboratory on Tuberculosis, Beijing Key Laboratory for Drug Resistant Tuberculosis Research, Beijing Chest Hospital, Capital Medical University, Beijing Tuberculosis and Thoracic Tumor Institute, Beijing, 100149, China; Beijing Key Laboratory of Drug Resistance Tuberculosis Research, Department of Pharmacology, Beijing Tuberculosis and Thoracic Tumor Research Institute, Beijing Chest Hospital, Capital Medical University, Beijing, 100149, China; University of Cologne, Department of Internal Medicine, Division of Infectious Diseases, 50931 Cologne, Germany; German Center for Infection Research (DZIF), Bonn-Cologne, Germany

## Abstract

Evidence has emerged that some healthy individuals with a history of exposure to *Mycobacterium tuberculosis* (Mtb) can develop protective antibody responses. However, it is not known whether patients with active tuberculosis elicit protective antibodies, and if they do, which bacterial antigens are targeted. To investigate the B cell responses during active infection, we generated a panel of monoclonal antibodies isolated from memory B cells of one patient. The antibodies, members of four distinct B cell clones, were directed against the Mtb phosphate transporter subunit PstS1. Antibodies p4–36 and p4–163 from two different B cell clones showed protective efficacy against Mtb and *Mycobacterium bovis-*BCG in an *ex vivo* human whole blood growth inhibition assay. Germline versions of p4–36 and p4–163 could no longer bind Mtb, implying that affinity maturation was vital for their activity. Crystal structures of p4–36 and a closely related clonal variant of p4–163, p4–170, complexed to PstS1 were determined at a resolution of 2.1Å and 2.4Å and revealed that the two antibodies recognize two distinctive epitopes on PstS1. As a proof of principle, p4–36 and p4–163 were used in a passive vaccination setting in aerosol Mtb-infected Balb/c mice, where both antibodies reduced bacterial lung burden by 50% after a single injection prior to Mtb infection. Our study shows that inhibitory B cell responses arise during active tuberculosis and identifies PstS1 as a target for elicitation of anti-Mtb antibodies.

## Introduction

Exposure to *Mycobacterium tuberculosis* (Mtb) results in a spectrum of outcomes, one of which results in active tuberculosis (ATB) disease^1^. Both innate and adaptive arms of the immune response have been implicated in immunity to Mtb^2,3^, but the role of humoral immunity and more specifically, antibodies, remains controversial^4,5^. Several studies have suggested that antibodies may play a protective role in at least a proportion of otherwise healthy individuals who have a history of exposure to Mtb^4,6–8^, and antibody responses have been correlated with protective efficacy of an experimental TB vaccine^9^. During active disease, higher antibody titers to the mycobacterial glycolipid lipoarabinomannan (LAM) have been correlated with decreased severity of infection^10^, and there is evidence of B cell dysfunction during active disease that resolves following treatment^11^. However, functional characterization of monoclonal antibodies isolated from patients with ATB have not been carried out, and it is not clear whether such antibodies exhibit any anti-bacterial effects.

In the present study, we explored B cell responses during ATB, while focusing on PstS1, a subunit of the Mtb phosphate transporter and a dominant antigen during infection^12,13^. We isolated and analyzed 85 monoclonal antibodies (mAbs) from one patient with elevated anti-PstS1 responses and found two antibodies p4–36 and p4–163 from distinct clones that demonstrated inhibitory activity against both BCG and Mtb. Structural analysis revealed that the two antibodies are directed against two different epitopes on PstS1 and do not compete with one another. When administered prior to infection of Balb/c mice with Mtb, both antibodies caused a modest reduction in lung bacterial burden, supporting that both p4–36 and p4–163 exhibit an anti-bacterial activity. Our study provides a proof of concept that protective antibody responses can be generated during the course of active tuberculosis disease and may inform both therapeutic and preventative vaccine design.

## Results

### Isolation of PstS1-specific mAbs

We recruited 26 in-patients with ATB (Extended Data, Table 1). All patients received standard treatment, recovered, and were eventually discharged. We screened patient sera for anti-Mtb antibody responses prior to initiation of antibiotics using lysates from two pathogenic Mtb strains, H37Rv and CDC1551. Unlike healthy community controls, the majority of patients (23/26) showed serum reactivity by ELISA to at least one lysate (Fig. 1a and 1b). Testing of antibody titres against soluble lysate versus cell-wall components of non-pathogenic Mtb-H37Ra^14^ revealed responses against both components, but with higher reactivity against the Mtb surface (Fig. 1c). To identify antibody reactivity against specific Mtb proteins, we tested sera against five selected surface-exposed proteins (Extended Data Fig. 1) that had been described previously to elicit antibody responses in human infection (Fig. 1d-h)^15,16^. The strongest response was directed against PstS1^12^ (Fig. 1d), which has been implicated in Mtb virulence^13^ and is one of three genes previously identified to contain amino acid sequence variants undergoing diversifying selection in conserved epitopes^17,18^. Therefore, we decided to characterize antibody responses to PstS1.

**Figure 1:**
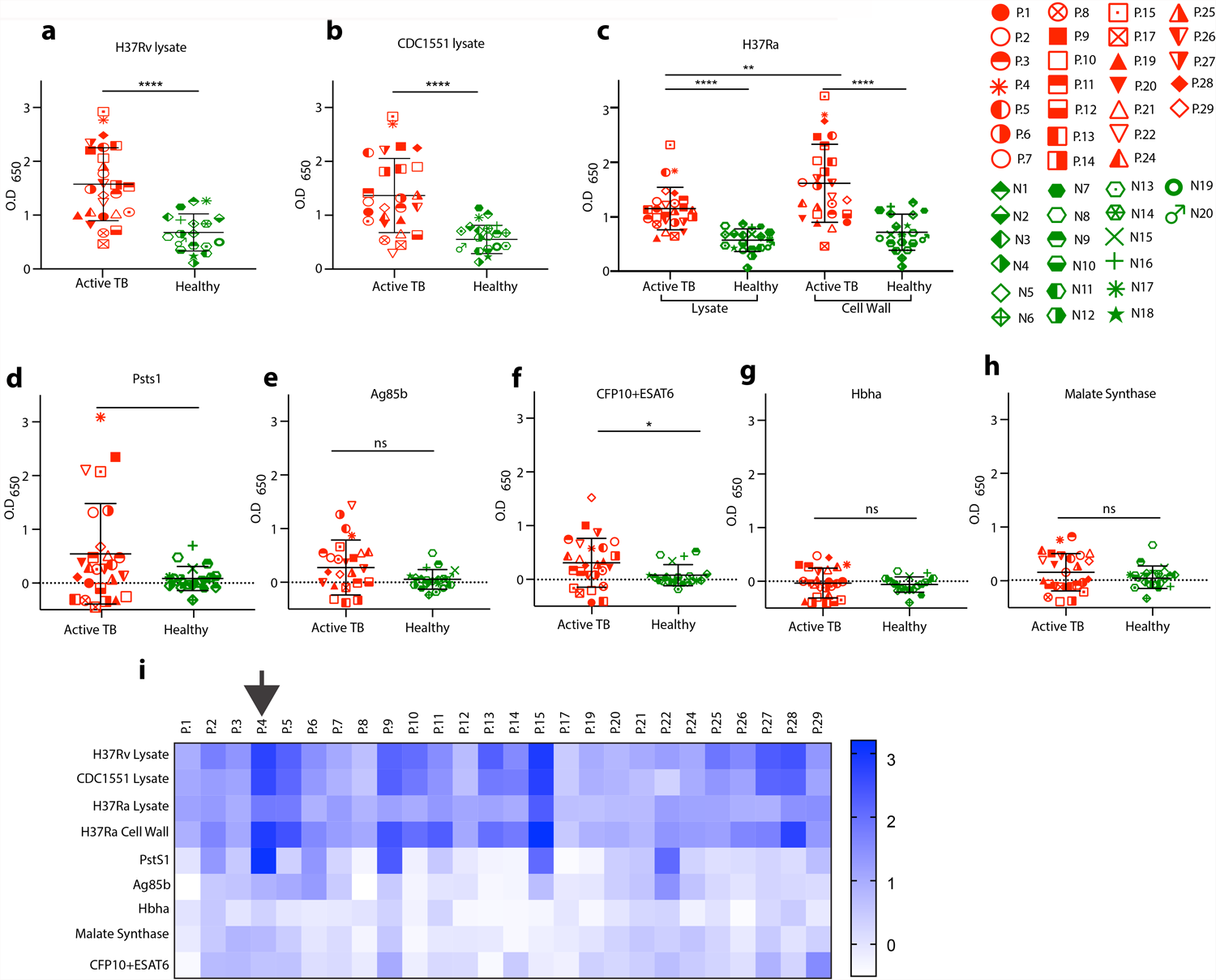
Serological profile of 26 actively infected tuberculosis patients. Serum responses by ELISA against **(a)** Mtb-H37Rv lysate, **(b)** Mtb-CDC1551 lysate, **(c)** Mtb-H37Ra lysate and cell-wall fraction, and recombinant proteins **(d)** PstS1, **(e)** Antigen 85b (Ag85b), **(f)** CFP10+ESAT6 complex, **(G)** Hbha, and **(h)** malate synthase (MS). Every symbol represents a single patient, with the legend given on the right side of the figure. ATB patients are in red symbols and negative community controls are in green. A heat map summarizing ELISA signals against the different antibodies and lysates is shown in **i**. Patient 004 is marked with a black arrow. Significance was determined by Welch’s t-test. *, p < 0.05, **, p < 0.01, ****, p < 0.0001.

We focused on one patient, P004, who had the strongest individual response to PstS1 and retained high anti-PstS1 responses over his treatment course (Fig. 1i and Fig. 2a). Hence, we sought to isolate monoclonal antibodies (mAbs) directed against PstS1 and test their effect on experimental models of Mtb infection. We isolated peripheral blood mononuclear cells (PBMC) from whole blood, and sorted single IgG+, PstS1+ B cells, which comprised 0.5% of the total IgG+ B cell population (Fig. 2b)^19^. A total of 102 heavy and 90 light chains were amplified by single-cell Ig PCR^20^; of these, 85 constituted natural heavy and light pairs (Extended Data, Fig. 2). Consistent with previous reports of anti-Mtb B cell responses in humans^7^, the majority of the antibody sequences were not clonal. Based on V_H_ D_H_ J_H_, V_L_, and J_L_ and > 75% identity in CDRH3^21^, a total of five clonal families were identified in 16 sequences (Fig. 2c). As previously described^21^, the sequences of the clonal heavy chains were more mutated when compared to the V_H_ sequences that were not part of a clone (Fig. 2d), but both groups had a similar V_H_ gene usage distribution (Extended Data, Fig. 2). We produced nine antibodies derived from four clones expressed in an IgG1 expression vector^20^ (Fig. 2e and 2f, expressed antibodies in red). We were unable to amplify the light chains from Clone 5 variants and therefore antibodies from this clone were not produced. All tested mAbs, except for p4–31, reacted strongly both with PstS1 and with Mtb lysates by ELISA (Fig. 2g-i). The mAbs also showed staining to H37Ra-mCherry compared with isotype control (Extended Data, Fig. 2). Amongst the nine antibodies we produced, mAbs p4–36 and p4–163 exhibited the strongest binding to bacterial lysates. We therefore decided to focus on these antibodies for the rest of the study.

**Figure 2:**
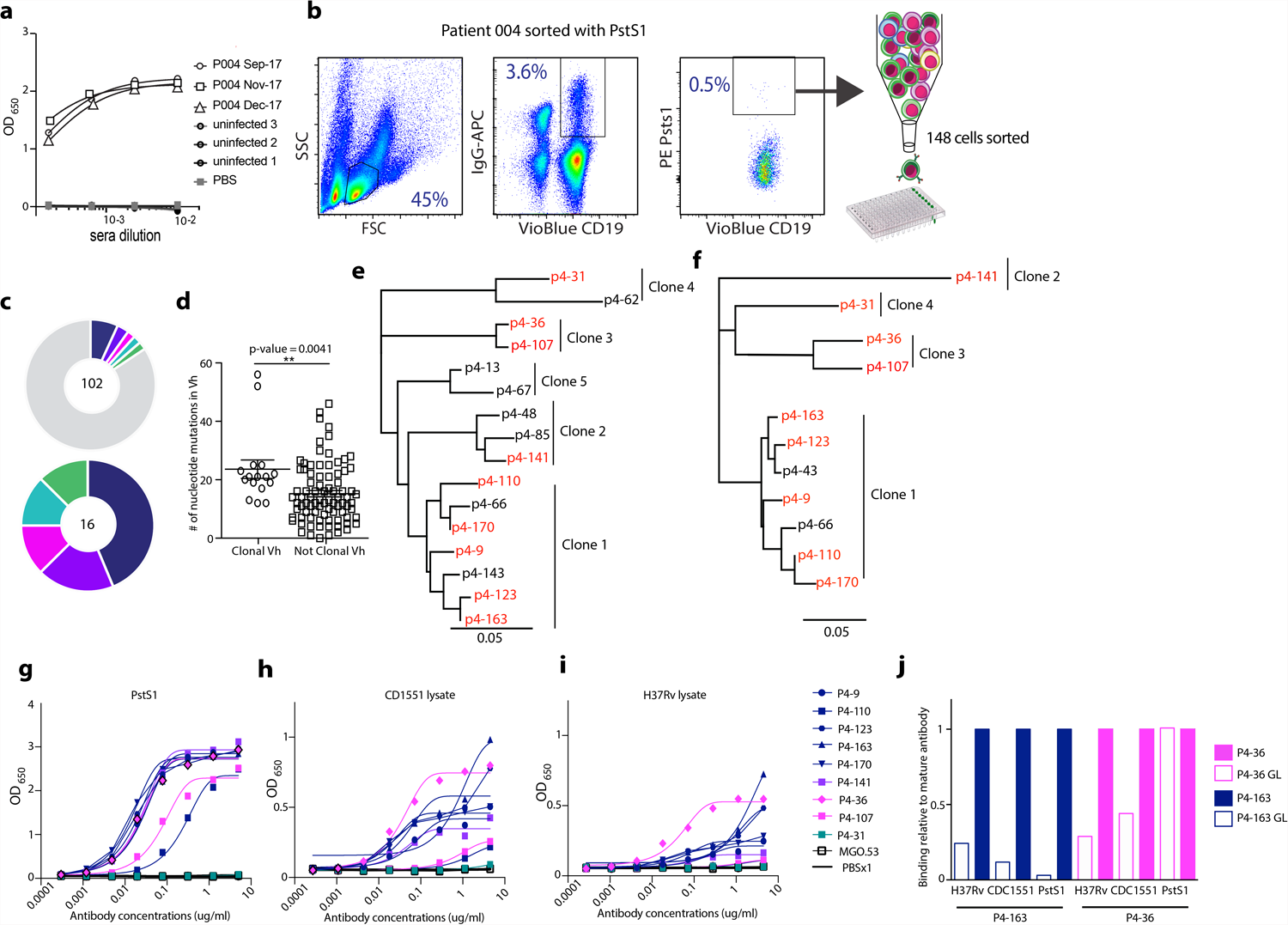
Isolation of anti-PstS1 antibodies from memory B cells of patient 004. **(a)** Patient 004 serum responses taken at three different time points to recombinant PstS1 by ELISA. **(b)** Whole blood-derived lymphocytes were stained for CD19, membrane IgG, and PstS1. A total of 148 positive cells were single-cell sorted. **(c**) Pie chart representing the heavy-chain sequences that were amplified from the CD19+/IgG+/PstS1+ B cells from patient 004. The number in the middle of the pie denotes the total number of sequences obtained, and the colored slices indicate clonally-related sequences. Upper panel: all sequences. Lower panel: only the 16 clonally related sequences. **(d)** Nucleotide mutations in V_H_ of the clonal versus non-clonal sequences. **(e)** and **(f)** are dendrograms of the clonally-related sequences, heavy and light chains, respectively, while the mAbs selected for expression are colored in red. The binding of 9 mAbs by ELISA to recombinant PstS1 **(g)**, Mtb-CDC1551 lysate **(h)**, and Mtb-H37Rv lysate **(i). (j)** Comparing binding by ELISA of the mature (mt) and the predicted germline (gl) versions of p4–36 (magenta) and p4–163 (dark blue), to recombinant PstS1, Mtb-CDC1551 lysate, and Mtb-H37Rv lysate. The binding score was determined as relative to the mature antibody, that was normalized to 1.

First, since the anti-PstS1 mAbs were generated during natural infection occurring in patient P004, we sought to investigate the impact of affinity maturation on their activity. Both p4–36 and p4–163 showed relatively low levels of somatic hypermutations, with p4–36 exhibiting 6 and 10 amino acid changes in heavy and light chains, respectively, and p4–163 exhibiting 10 and 9 amino acid changes in heavy and light chains, respectively. Nevertheless, we hypothesized that these mutations provide the mAbs with their ability to bind PstS1 and Mtb lysates. To test our hypothesis, we reverted the amino acid sequences of p4–36 and p4–163 back to their germline sequences similarly to what was previously carried out for other naturally elicited mAbs (Extended Data, Fig. 4)^22^, thus producing the un-mutated predicted germline (gl) versions of each antibody: p4–36gl and p4–163gl. The binding of the germline variants was compared to the binding of the mature antibodies to PstS1, as well as H37Rv and CDC1551 Mtb lysates. Both mAbs showed 2- to 10-fold reduction in their binding when somatic hypermutations were reverted, with the exception of mAb p4–36gl that bound recombinant PstS1 with similar affinity as the mature p4–36 (p4–36mt), but not the bacteria (Fig. 2j). To further investigate whether mutations occurring in the heavy chain or the light contribute to antibody activity, we produced chimeric antibodies where the heavy chain is mature while the light chain is germline (HCmtLCgl), and *vice versa* (HCglLCmt). Testing those for binding showed that for p4–36 mutations in the light chain were more significant for binding than those in the heavy chain, as opposed to p4–163 where mutation in the heavy chain were more essential for binding compared to mutations in the light chain (Extended Data Fig 4 c-e). Overall, we conclude that both mAbs p4–36 and p4–163 are specific for PstS1 and acquired somatic hypermutations during natural infection that resulted in their improved binding to Mtb.

### Activity *ex vivo*

Next, we sought to test the effects of the mAbs p4–36 and p4–163 on Mtb infection in culture. First, we tested the effect of our mAbs on bacterial entry. Infecting phorbol 12-myristate 13-acetate-(PMA)-differentiated THP-1 macrophages^23^ with the attenuated H37Ra strain with and without PstS1-specific mAbs. The mAbs bound H37Ra within macrophages and did not prevent, but rather slightly increased, bacterial entry into the macrophages (Fig. 3a). We next asked whether the mAbs p4–36 and p4–163 could inhibit mycobacterial growth. For this, we used a whole-blood mycobacterial growth inhibition *ex vivo* assay (MGIA) where human PBMCs from healthy donors are infected with pathogenic bacteria^24^. This system has been used previously to test polyclonal antibodies from healthy donors, and allows testing of antibody activity in a more physiologically relevant *in vitro* system. Here, p4–36 and p4–163 were able to significantly restrict growth of both *M. bovis*-BCG (BCG) and pathogenic Mtb (Fig. 3b,c).^25^,^24^. This activity was not dose dependent and was unique to mAbs p4–36 and p4–163, and was not observed with other anti-PstS1 mAbs, such as p4–31 and p4–141, showing that ELISA binding to Mtb lysates correlated well with their activity *ex vivo*.

**Figure 3:**
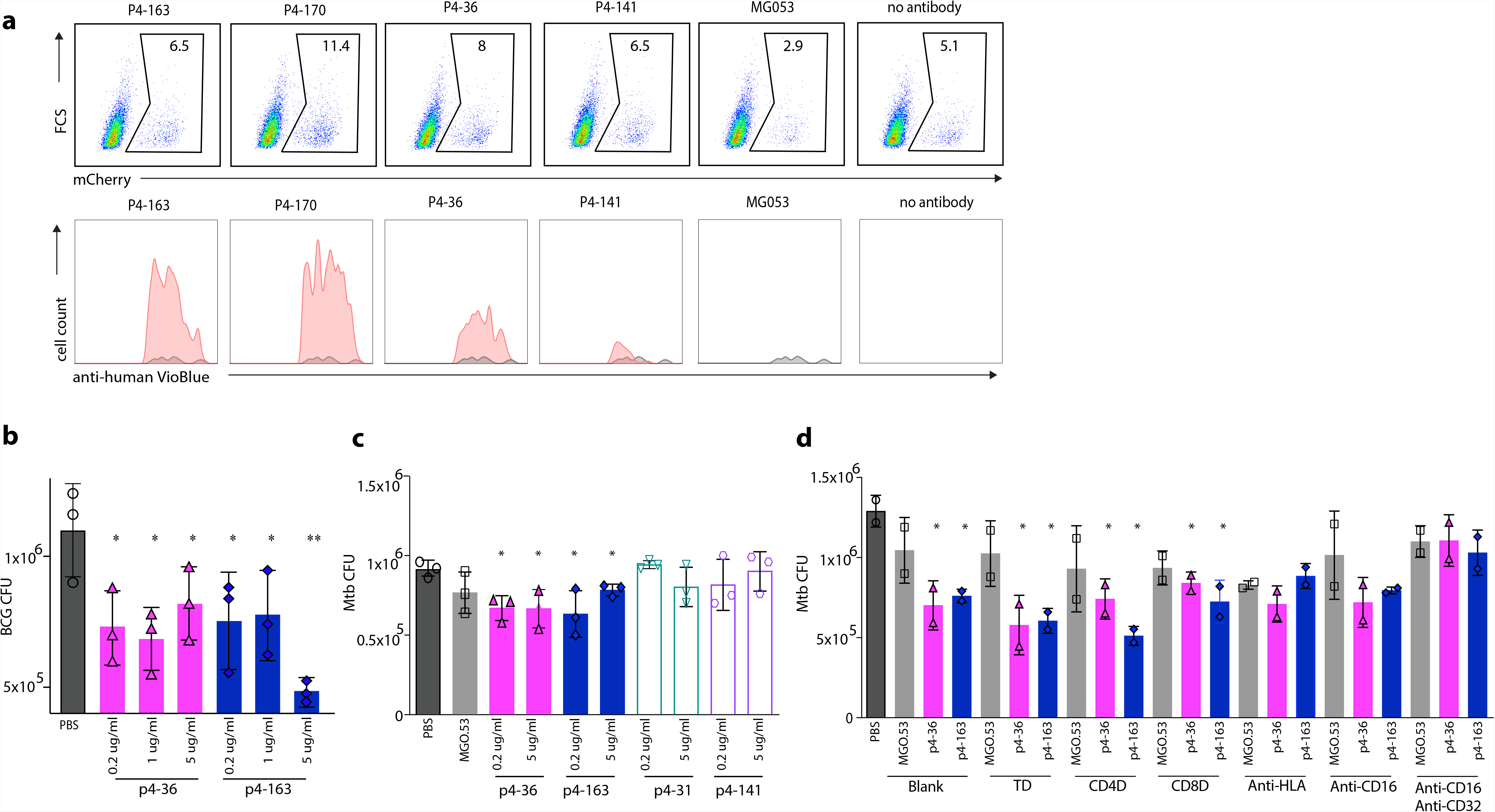
Anti-PstS1 mAbs inhibit Mtb in culture. **(a)** Upper panel: gating strategy of H37Ra-infected, mCherry positive, PMA-differentiated THP-1 cells pre-incubated with mAbs p4–163, p4–170, p4–36, p4–141, and the isotype control mAb MGO.53^20^ by flow cytometry. Lower panel: histograms showing the intensity of signal and cell counts of mAb-bound intracellular bacteria as indicated by anti-human Vioblue staining. For each mAb, the binding histogram is shown in red and compared to the isotype control histogram, which is depicted in a gray overlay. **(c)** and **(d)** Activity of anti-PstS1 mAbs at indicated concentrations in a human whole-blood mycobacterial growth inhibition assay (MGIA) after 96 hours of infection with BCG or pathogenic Mtb, respectively. **(d)** Activity of anti-PstS1 mAbs (5 µg/ml) in MGIA following depletion of CD3+ T cells (TD), CD4+ T cells (CD4D), CD8+ T cells (CD8D), blockade of MHC II (anti-HLA), CD16 or CD32 or both (anti-CD16, anti-CD32); see Methods. Significance was determined by Student’s t-test.*, p < 0.05, **, p < 0.01.

We next sought to investigate the mechanism of mAb-induced CFU reduction over time. MAbs p4–36 and p4–163 did not inhibit infection, and immediately after infection more bacteria were found in THP-1 cells (Fig. 3a). We suspected an antibody-dependent opsonization of bacteria *via* Fc gamma receptors^6,8,24^. To test this, we repeated the MGIA experiments with Fc receptor blockers. Blocking the main Fc gamma receptors on human macrophages, CD16 and CD32, eliminated the inhibitory activity of both p4–36 and p4–163 (Fig. 3d). By contrast, specific depletion of T cells did not affect mAb-induced CFU reduction (Fig. 3d). We conclude that p4–36 and p4–163 reduce Mtb CFU over time in an Fc-dependent manner. Whether this Mtb inhibition is due to an active restriction of intracellular bacterial replication or due to induction of bacterial killing is yet to be determined.

### Structure-function studies

To further understand the mechanism of the protective antibody binding to its target, we produced antigen binding fragments (Fabs) of p4–163 and p4–36 and prepared Fab-PstS1 complexes for crystallization. We determined the structure of Fab p4–36 in complex with PstS1 at a resolution of 2.1 A (Fig. 4a and Extended Data, Table 2). Two PstS1-Fab p4–36 heterodimers were in the asymmetric unit of the crystal and had only minor differences in the constant domains of the bound Fabs (Extended Data, Fig. 5). P4–36 binds a contiguous epitope located on an alpha-helix formed by residues 141–145 and preceding residues 136–137 and residues 139–140 (Fig. 4b and Extended Data, Fig. 6a), within a small surface area of 583 A□^2^. The contacts between p4–36 and PstS1 are mostly Van der Waals and hydrogen bonds, and contributed largely by complementarity determining regions 1 (CDR1) and CDR3 of the light chain (CDRL1 and L3) and CDR3 of the heavy chain (CDRH3) (Fig. 4b and Extended Data, Fig. 7a and 7b and Extended Data Table 3). Ten hydrogen bonds are observed at the interface, and one salt bridge is formed between Asp36^p4–36 CDRL3^ and Lys136^PstS1^ (D36^L^[OD1]-K136^PstS1^[NZ]) (Fig. 4b).

**Figure 4:**
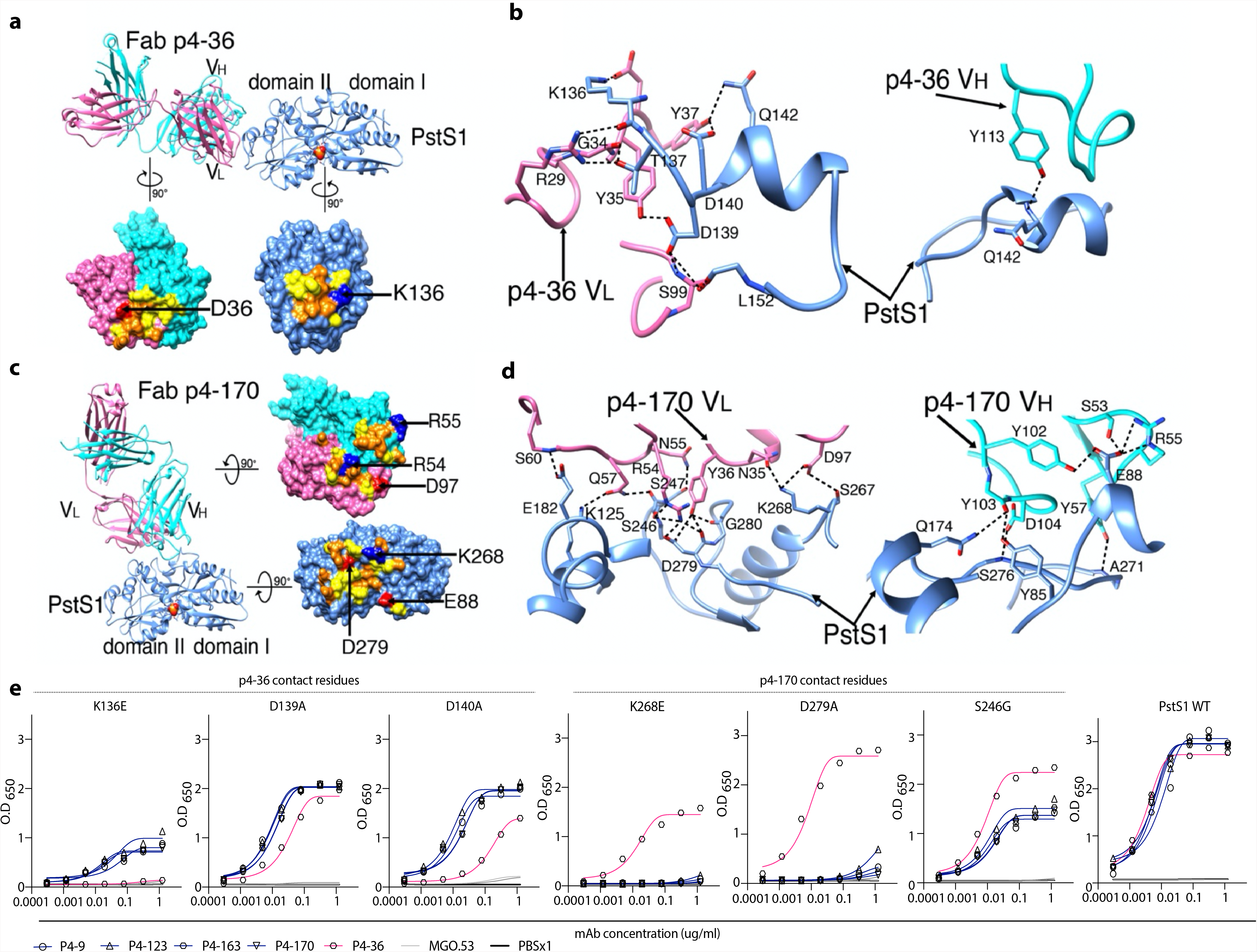
MAbs p4–170 and p4–36 recognize different epitopes on top of PstS1. **(a)** Top: Ribbon diagrams show the crystal structure of PstS1 in a complex with Fab p4–36. The Fab p4–36 heavy chain and light chain are colored cornflower cyan and hot pink, respectively. The PstS1 structure is colored blue. The bound phosphate (Pi) is represented as filled balls with oxygen and phosphorus atoms colored red and yellow, respectively. Bottom: An open-up surface-shadowed representation showing the contact interface. Residues involved in hydrogen bonds and Van der Waals contacts are highlighted in orange and yellow, respectively. Positively and negatively charged residues involved in the formation of the salt bridges are highlighted in blue and red, respectively. **(b)** Close-up view of the interface between PstS1 and Fab p4–36. The dashed lines indicate hydrogen bonds and salt bridges. **(c**) Left: Ribbon diagrams show the crystal structure of PstS1 in complex with Fab p4–170. Right: An open-up surface-shadowed representation showing the contact interface. The color schemes used are the same as in (a). **(d)** Close-up view of the interface between PstS1 and Fab p4–170. The dashed lines indicate hydrogen bonds and salt bridges. **(e)** Four Clone 1 variants (mAbs p4-9, p4–123, p4–163 and p4–170, and mAb p4–36), as well as negative control mAb MGO.53, were tested for binding by ELISA to six point mutant PstS1 proteins. The upper panel shows binding curves of PstS1 mutated in p4–170 contact residues K268E, D279A, and S246G, while the lower panel depicts binding curves of PstS1-mutated p4–36 contact residues K136E, D139A, and D140A. The legend is given at the bottom of the figure, and the binding curves to wild type PstS1 are shown in the right panel of the figure.

No crystal could be obtained with the complex of Fab p4–163 and PstS1. Thus, we switched to p4–170, which is a closely related variant of p4–163 and a member of the same B cell clone (Clone 1, Fig. 2e,f), and has 97.6% and 93.86% amino acid sequence identity to p4–163 heavy and light chain, respectively. The Fab p4–170 and PstS1 complex could be crystallized, and the structure was determined at a resolution of 2.4 A□ with one PstS1-Fab p4–170 heterodimer in the asymmetric unit (Fig. 4c and Extended Data, Table 2). As revealed by the structure of the complexes, p4–170 binds to a different epitope from the one recognized by p4–36.

The epitope recognized by p4–170 is highly discontinuous and conformationally proximate to the Pi binding site in the middle of the molecule, between the two domains I and II of PstS1 (Fig. 4c). The epitope is a large surface of 977 A□^2^ and is comprised of three helices and four loops of PstS1 (Fig. 4d and Extended Data, Fig. 6b). The interactions between p4–170 and PstS1 are mediated through salt bridges, hydrogen bonds, and Van der Waals contacts (Fig. 4d and Extended Data, Table 4). Except for CDRH1, all the CDRs of p4–170 are involved in the interactions with residues 85, 88, and 92 of the PstS1 domain I and residues 125, 174, 177–178, 181–182,190, 246–248, 267–268, 270–271, 275–276, and 279–281 of domain II (Fig. 4d and Extended Data, Fig. 7c and 7d and Extended Data Table 4). The hydrogen bonds are formed with both the main chain atoms and side chain atoms of PstS1. Four salt bridges are formed at the contact interface, including Glu88^PstS1^[OE1]-Arg55^H^[NE], GLu88^PstS1^[OE1]-Arg55^H^[NH2], Lys268^PstS1^[NZ]-Asp97^L^[OD1], and Asp279^PstS1^[OD2]-Arg54^L^[NH2] (Fig. 4c and Extended Data, Table 4).

To confirm the key antibody:antigen contact residues identified by the structures, we generated a panel of point mutations in PstS1. Mutating residues at the p4–170:PstS1 interface S246G^PstS1^, K268E^PstS1^, and D279A^PstS1^ reduced the binding of all tested Clone 1 mAb variants, but not the binding of p4–36 (Fig. 4e). Amino acid substitutions in the alpha-helix that holds the epitope of p4–36 residues K136E^PstS1^, D139A^PstS1^ and D140A^PstS1^ completely abolished p4–36 binding. The binding of Clone 1 mAbs was also reduced, indicating that the alpha-helix bound by p4–36 might be essential for the folding of PstS1.

We next asked whether the binding of p4–36 of p4–170 could interfere with multimerization of the transporter. For this, we performed modeling the binding of PstS1 to the transporter complex. According to our prediction, the bound antibodies show no blockage to the assembly of the PstA-B-C-S complex and do not block the binding of PstS1 to the PstA, B, C transporter complex. We conclude that p4–36 and p4–170 does not function by inhibiting the transporter activities (Fig. 5).

**Figure 5:**
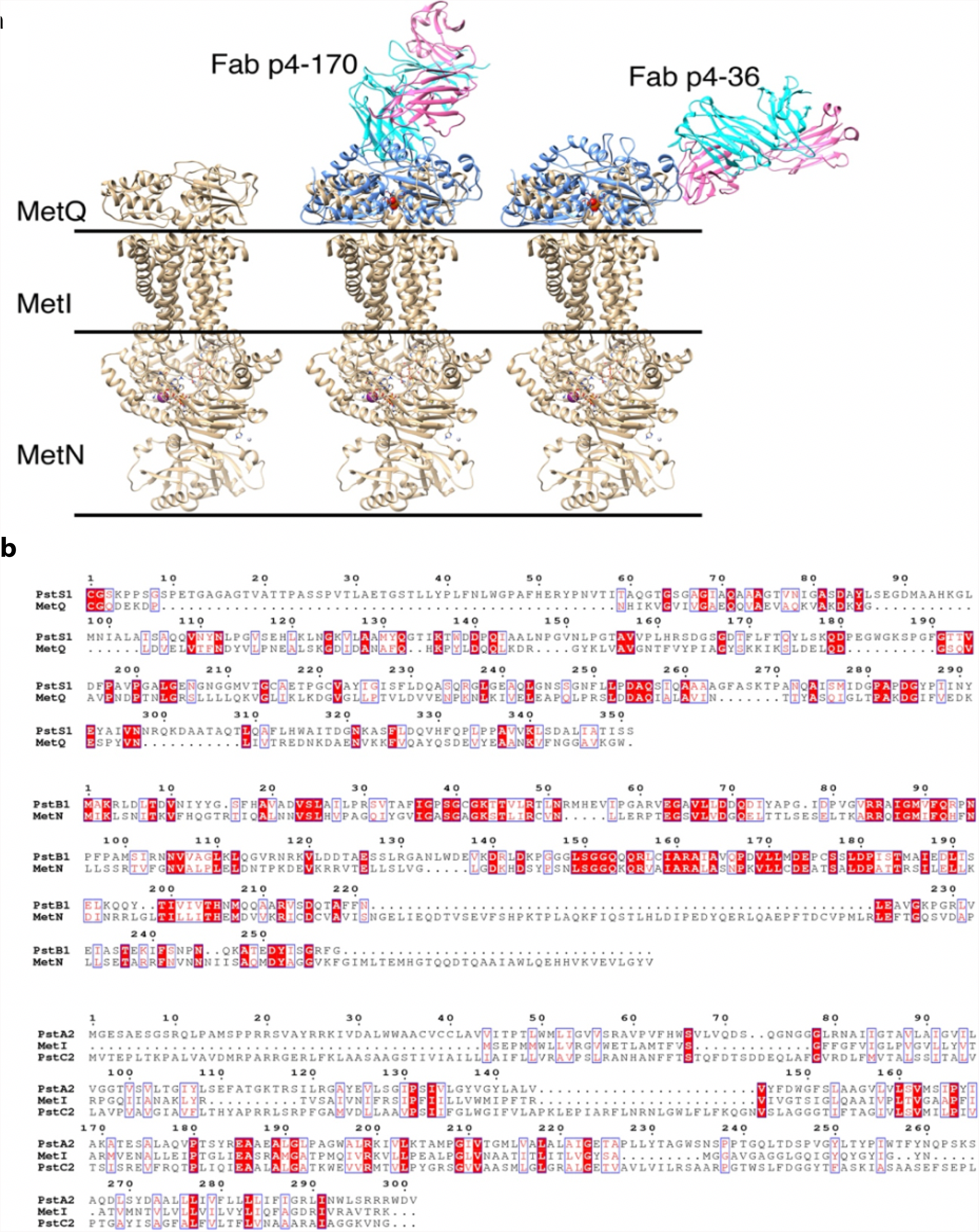
Structure modeling of the binding of PstS1 to the PstA-B-C phosphate transporter complex. **(a)** Structural superimposition of the Fab – PstS1 complexes with MetQ of the MetNIQ ABC transporter complex (PDB code: 6CVL) suggested a possible binding mode of PstS1 to the PstA-B-C phosphate transporter complex. The bound antibodies show no blockage to the assembly of the PstA-B-C-S complex. **(b)** Sequence alignments showing that the MetNIQ complex components share significant sequence similarities to these of the PstA-B-C phosphate transporter complex of Mtb.

### Activity *in vivo*

Finally, we tested the activity of the two mAbs p4–36 and p4–163 on Mtb infection *in vivo*. For this, we used wild type Balb/c mice. We injected 0.5 or 1 mg mAb per mouse intra-peritoneally five hours prior to aerosol infection with pathogenic Mtb. After two weeks the mice were sacrificed and lung bacterial burden determined. Lung burdens were reduced in mice pretreated with mAbs p4–36 and p4–163, or both by approximately 0.5 log CFU (Fig. 6), which agrees with the MGIA results and verifying that the mAbs have anti-Mtb inhibiting activity.

**Figure 6:**
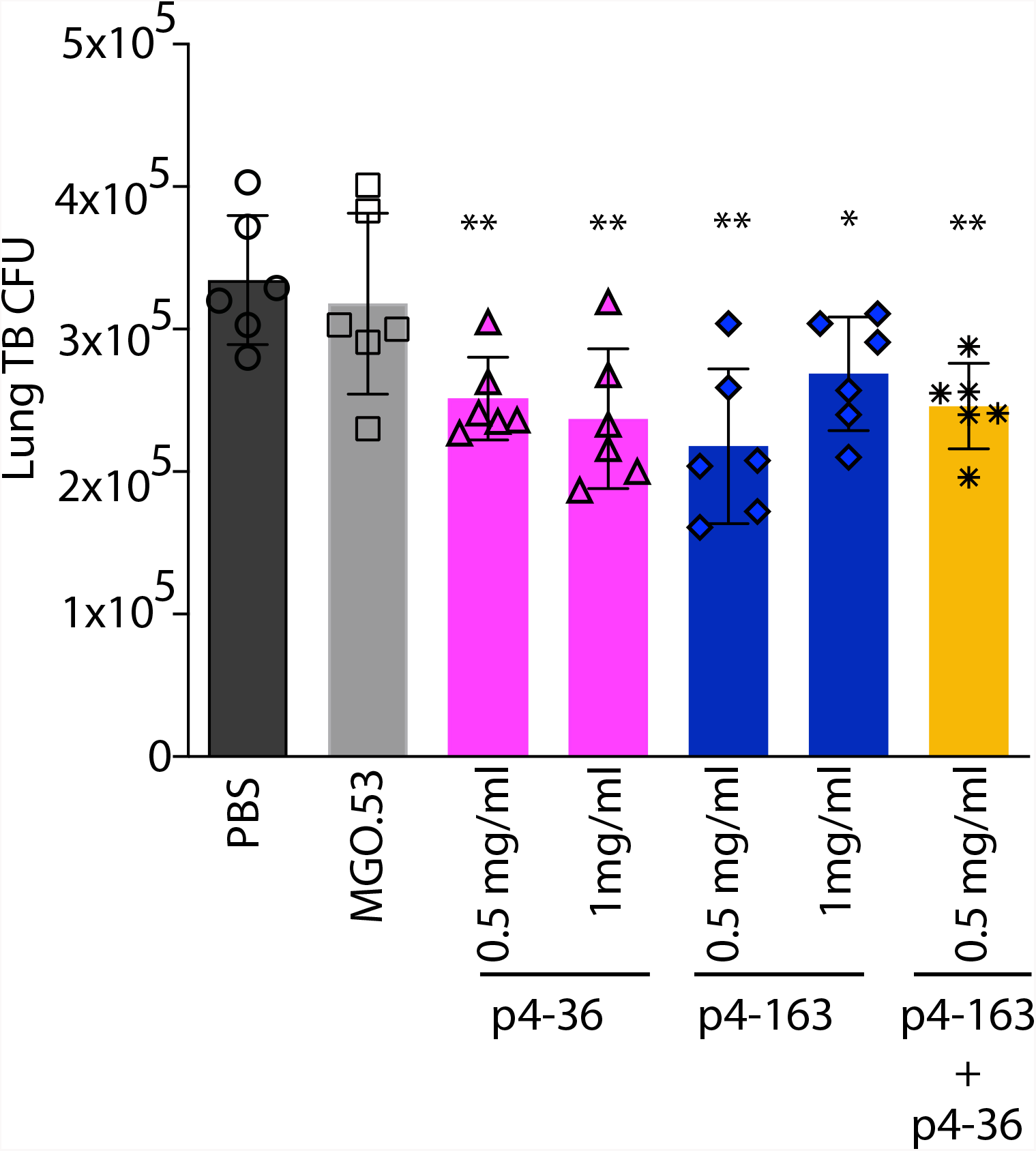
Anti-PstS1 mAbs inhibit Mtb in Balb/c mice. Activity of anti-PstS1 mAbs p4–36 (magenta) and p4–163 (dark blue) at indicated concentrations in Balb/c mice. Mice were injected once intra-peritoneally with mAbs at indicated amounts 5 hours prior to aerosol infection with Mtb. Mice were sacrificed after two weeks, and lung bacterial burden measured. Results are representative of two independent experiments. Significance was determined by Student’s t-test. ns, no significance,

## Discussion

Here, we describe a patient, P004, who had active tuberculosis and was selected from a cohort of infected patients due to his strong anti-Mtb serum responses to the PstS1 antigen. While the activity of the two anti-PstS1 mAbs we isolated is modest (about 0.5 Log), to our knowledge we are the first ones to describe PstS1 as a potential target for naturally-elicited human antibodies that have anti-Mtb activity.

PstS1 is a 38-kDa phosphate-binding periplasmatic protein, one of three subunits of the Mtb phosphate-specific transporter (Pst) complex^12^. PstS1 is as an immunodominant marker for ATB^26–28^. PstS1 is also necessary for Mtb virulence; PstS1-deletion mutants were attenuated in a mouse-infection model^13^. Intriguingly, PstS1 was identified as one of only three Mtb genes that are subject to evolutionary sequence diversification, suggesting that PstS1 variation plays a role in Mtb immune evasion^17,18^.

In the present study we characterize two different sites on PstS1 that can be targeted by antibodies capable of inhibiting Mtb growth. P4–163 is a member of the largest B cell clone of 9 mAbs that shared > 93% sequence identity. The high-resolution structure of a clonal variant of p4-163, p4–170, revealed a large, sparse, and highly conformational epitope that is adjacent to the active site of PstS1. Intriguingly, seven of p4–170 contact residues (Lys268^PstS1^, Pro270^PstS1^, Ala271^PstS1^, Ile275^PstS1^ Ser276^PstS1^, Asp279^PstS1^, and GLy280^PstS1^) overlap with a highly conserved Mtb T cell epitope 259-AAAGFASKTPANQAISMIDG-280, Immune Epitope Database number 35^17,29^. The epitope of p4–36, a member of a different and smaller B cell clone, bound to a condensed alpha-helix structure within residues 136^PstS1^-145^PstS1^, located 30 A distant from the epitope bound by p4–170.

P004 had active tuberculosis; therefore, by definition, the mAbs we isolated did not prevent or eliminate Mtb infection in this patient. However, evidence from other infectious diseases shows that B cell responses develop in parallel and subsequent to high pathogen loads that stimulate B cell activation and maturation, causing neutralizing/protective antibodies to arrive ‘too little, too late,’ without a significant benefit to the individual that produced them^22,30,31^. Considering the low extent of affinity maturation, as reflected by the relatively low number of somatic hypermutations in p4–36 and p4–163, it is possible that these B cell clones only started their evolution and did not reach their full anti-Mtb potential. On the other hand, the fact that these antibodies ultimately arise provides evidence that B cells play an active role during ATB.

Despite long-standing controversy, recent studies have demonstrated that in certain cases anti-Mtb humoral responses can be protective^7,24,32^ or correlate with lack of active disease or even lack of infection^4,6,32,33^. Only one prior study profiled the human B cell responses against Mtb antigens on a monoclonal level^7^. This study attributed bacterial inhibition to IgA isotypes rather than IgG. However, in that study, ‘protection’ was defined as decreased viable Mtb within cells shortly after infection. Unfortunately, the study evaluated neither *in vivo* activity nor the precise molecular mode of antigen binding. While Balb/c mouse model used in this study does not recapitulate lung granulomas which are the hallmark of active TB infection in humans, passive transfer of mAbs before pathogenic Mtb infection resulted in significant reduction of bacterial lung loads in mice indicating a preventive mAb activity. This is the first report of human anti-Mtb mAbs that have been shown to have a modest, yet robust, activity *in vivo*, as well as resolving their structure along with their corresponding Mtb target. Our study has implications for the development of new anti-Mtb therapies. Additionally, the fact that the germline version of p4–36, p436gl, bound recombinant PstS1 suggests that PstS1 could be used as an immunogen to elicit p4–36-like antibodies in naïve populations.

## Materials and Methods

### Study design

The objective of this study was to investigate whether antibodies isolated during Active Tuberculosis (ATB) disease can inhibit Mtb *in vivo* and elucidate their corresponding Mtb target and mechanism of action. Donor 004 was selected based on exceptional serum binding to Mtb lysates and to recombinant PstS1 protein. Staining with PstS1 and single-cell sorting of antigen-positive memory B cells from donor 004 allowed isolation of four new clones of antibodies, two of which were able to inhibit the growth of pathogenic Mtb *ex vivo* and *in vivo*. We analyzed structurally the two mAbs and discovered that they are directed against two non-overlapping sites on PstS1. Their ability to inhibit Mtb growth was tested by infecting healthy human PBMCs and in vivo, in Balb/c mice. The epitopes were identified by x-ray crystallography.

### Ethics statement

For the patient studies, the Tel Aviv University Institutional Review Board (IRB) approved all studies involving patient enrollment, sample collection, and clinical follow-up. Donor 004 provided written informed consent prior to participating in this study, and The Tel Aviv University and Shmuel Harofe Hosptial Institutional Review Boards approved all studies involving patient enrollment, sample collection, and clinical follow-up (protocols number 33.18. and 0058). Donor P004 was selected from a group of twenty-six active pulmonary TB patients that were followed by Pulmonary and Tuberculosis Department of Shmuel Harofe hospital in Israel, and is also referred to as subject ID 109004. Fresh human whole blood for MGIA assays was obtained from volunteers under IRB-approved protocol (2014–2–25) of the Beijing Tumour and Thoracic Hospital. For the mouse studies, this study was carried out in strict accordance with the guidelines of the Chinese Association for Laboratory Animal Sciences, and approved by the institutional ethical Committee (IEC) of Peking Union Medical College.

### Study participants

Twenty-six active pulmonary TB patients were recruited from the Pulmonary and Tuberculosis Department of Shmuel Harofe hospital in Israel. Patients were diagnosed with active pulmonary TB by sputum smear and positive bacterial culture, and treated with antibiotics. Blood samples for screening for anti-Mtb serum antibodies were drawn before receiving antibiotic therapy. For patients who exhibited high anti-Mtb serum activity, blood draw was repeated two additional times during hospitalization. Patient 004, ID 109004 (Extended data, Table 1), a 35-year-old male, was diagnosed with drug-sensitive TB, and a small blood donation was first collected on September 9^th^, 2017. This donor was asked for a large donation of whole blood, from which peripheral blood mononuclear cells (PBMCs) were isolated by Ficoll gradient density centrifugation. Serum samples of healthy donors served as controls and were obtained from the Israeli blood bank. TB exposure history of these subjects was not available.

### Bacterial culture, lysate, and cell wall fractions

Lysates of Mtb-H37Rv (Cat# NR-14822) and the Mtb-CDC1551 (Cat # NR-14823) were obtained from BEI resources https://www.beiresources.org/. For the preparation of H37Ra^14^ lysate and cell wall fractions, H37Ra bacterial culture was grown in Middlebrook 7H9 broth supplemented with 10% OADC, 0.05% Tween-80, 0.5 % glycerol, at 37 °C in a shaking incubator to reach an optimal density of O.D_600_ = 0.4. The bacterial culture was centrifuged at 4,000 rpm for 15 min at room temperature. The pellet was resuspended in PBSx1 containing 1% PMSF (Sigma) and was lysed by sonication. Following centrifugation at 14,000 rpm for 30 min at 4 °C, the lysate was clarified, and the cell wall fraction was resuspended in PBSx1.

### Expression of recombinant Mtb antigens

Mtb antigens PtsS1 (Rv0934), Ag85b (Rv1886c), Hbha (Rv0475), CFP10 (Rv3874), ESAT6 (Rv3875), and Malate Synthase (Rv1837) were cloned into a pMALp vector as fusion to maltose binding protein (MBP), with a TEV cleavage site separating the recombinant Mtb proteins and MBP (Extended Data, Fig. 1). The recombinant Mtb proteins were C-terminally tagged with 6xHis and a specific BirA biotinylation sequence and were expressed in *E-coli* (BL21 strain). Shortly, plasmid-transformed *E-coli* were grown to an optimal density of O.D_600_ = 0.6., at which, protein expression was induced by adding Isopropyl-β-D-1-Thiogalactopyranside (IPTG) (0.5 mM), and the culture was grown for 4 hours at 30 °C in a shaking incubator of 225 rpm. Next, bacteria were pelleted by centrifugation at 8,000 rpm for 15 min at 4 °C, and the pellet was frozen and kept at a temperature of-20 °C. Afterwards, bacterial pellets were thawed and resuspended in 50 mM NaH2PO4, 300 mM NaCl, 10 mM imidazole pH = 8, TritonX 0.1 % (Sigma), and protease inhibitor cocktail (Sigma) (“binding buffer”), and lysed by sonication. Following additional centrifugation at 14,000 rpm for 30 min at 4 °C, the protein was purified from the supernatant phase using Ni sepharose beads (GE Healthcare), TEV cleavage, and elution with 50 mM and 100 mM imidazole (Sigma). Enzymatic site-specific biotinylation was carried out by BirA biotinilation kit (Avidity). All recombinant Mtb proteins were stored in PBSx1.

### ELISAs

High-binding 96-well ELISA plates (Corning, 9018) were used for all experiments. For sera binding to Mtb strains, plates were coated overnight at 4 °C with 1–2.5 μg/ml of pathogenic bacterial lysates, H37Rv, and CDC1551 (both received from BEI recorces), or H37Ra lysate or cell wall fractions. For sera binding to recombinant Mtb antigens, plates were coated overnight at 4 °C with 5 μg/ml of PstS1, Ag85b, Hbha, Malate Synthase, and ESAT6+CFP10. ESAT6+CFP10 were premixed at a 1:1 ratio. The next day, after washing 3 times with PBS + 0.05% Tween-20, the plates were blocked for 2 hours at room temperature with 3% BSA, 20 mM EDTA, and 0.05% Tween-20 in PBS (“blocking buffer”), followed by a one-hour incubation at room temperature with polyclonal Mtb-infected sera or negative healthy controls at 1:100 and 1:300 dilutions in blocking buffer. After an additional washing step, the plates were incubated for 1 hour with peroxidase-conjugated goat anti-human IgG (Jackson ImmunoResearch) at a final concentration of 0.16 μg/ml at room temperature and developed by adding TMB substrate (abcam). The plates were read in an ELISA plate reader after 20 min at O.D_650_.

For anti-PstS1 and germline mAbs ELISA, plates were coated overnight at 4 °C with 5–10 μg/ml of recombinant PstS1 antigen or pathogenic bacterial fractions as described above. The next day, after washing and blocking as indicated above, plates were incubated for 1 hour at room temperature with anti-PstS1 or germline mAbs in eight consecutive 4-fold dilutions, starting from a concertation of 5 μg/ml. After an additional washing step, the plates were incubated with peroxidase-conjugated goat anti-human IgG, developed and read as indicated above.

For PstS1 mutants ELISA, plates were coated overnight at 4 °C with 5μg/ml of recombinant PstS1, wild type and mutants. The next day, after washing and blocking as indicated above, plates were incubated for 1 hour at room temperature with anti-PstS1 mAbs in eight consecutive 4-fold dilutions, starting from a concertation of 1.25 μg/ml. After an additional washing step, the plates were incubated with peroxidase-conjugated goat anti-human IgG, developed and read as indicated above.

### Single B cell sorting, sequencing, cloning, and expression of antibodies

Patients’ purified peripheral blood mononuclear cells (PBMCs) were isolated by ficoll (GE Healthcare) and frozen at liquid nitrogen. PBMCs were thawed and enriched for B cells by CD20 magnetic microbeads (MACS, Miltenyi Biotec). The CD20+ B cell fraction was stained for 15 min at 4 °C with anti-human antibodies: CD19-VioBlue (1:100; Miltenyi Biotec), IgG-APC (1:20; Miltenyi Biotec), and with biotinylated PstS1 pre-incubated with streptavidin-PE (1:10; Miltenyi Biotec). All the PstS1-PE positive IgG+ CD19+ cells were single-cell sorted into 4 ul of lysis buffer (PBS, RNAsin-Promega and 0.1 M DTT). Rescue primers were used to amplify both heavy chains^34^ and Igλ genes^35^, and regular primers were used for IgK chains^36^. All PCR products were sequenced and analyzed for Ig gene usage, CDR3, and the number of V_H_/V_L_ somatic hypermutations (IgBLAST, https://www.ncbi.nlm.nih.gov/igblast; and IMGT, https://www.imgt.org). Purified, digested PCR products were cloned into human Igγ1, Igk, or Igλ-expression vectors as previously described^36^ and produced by transient transfection of IgH, IgK, and IgL expression plasmids into exponentially growing Expi293F cells, as previously described^37^.

### Flow cytometry for direct and intracellular antibody binding

The binding of p4-mAbs to H37Ra-mCherry was determined as followed: Bacteria were grown to OD_600_ = 0.4, centrifuged, washed with PBS, and passed through a 30G syringe ten times for clump elimination. A total of 10^6^ colony forming units (CFU) were incubated with or without 50 μg/ml p4-mAbs for 1 hr at RT, washed with a FACS buffer (PBS, 1% FBS, 2mM EDTA), and incubated with anti-human IgG-VioBlue (1:50 dilution; Miltenyi Biotec) for 40 min at room temperature. The unbound antibodies were washed, and the bacteria were fixed and permeabilized (BioGems) and analyzed by cytoflex. For staining of intracellular THP-1-infected H37Ra-mCherry, human monocytes THP-1 cell line^23^ (ATCC) were cultured in RPMI medium (Biological Industries) and differentiated for 24 hours by addition of phorbol 12-myristate 13-acetate-(PMA) 160 ng/ml at 37 °C. On the day of infection, bacteria were incubated with 50 μg/ml p4-mAbs and MGO.53^20^, used as isotype control for 30 min at RT. The p4-mAbs and bacterial mix, along with anti-human IgG-VioBlue (1:200 dilution; Miltenyi Biotec), were added to 8×10^5^ differentiated THP-1 macrophages at MOI 1:10 for 3 hours at 37 °C. Following 5 washes with PBS, the cells were incubated with 250 μg/ml Amikacin sulfate salt (Sigma) for 1 hour at 37 °C. Cells were washed and dislodged using Cell Dissociation Solution (Biological Industries), and then fixed and permeabilized (BioGems).

### Mycobacterial Growth Inhibition Assay (MGIA)

Blood from healthy volunteer donors was drawn into a CPT vacutainer tube. Blood from the same donor was used for any given set of experiments, to rule out donor-mediated variability. The blood was aspirated into a new 50 ml Falcon tube, and sodium citrate (3.2g/L) was added as an anticoagulant at a ratio of 1:9 (sodium citrate/blood). Blood was diluted 1:1 with RPMI-1640 (Gibco). A local clinical isolate of pathogenic Mtb, Beijing strain (strain 165,^38^) was passed through a 5 µm filter to remove clumps. The OD was checked and diluted with the RPMI-1640 medium. Next, 0.1 ml of the bacterium (10^5^ CFU) was added to 0.9 ml of the diluted blood in 15 ml sterile falcon tubes. A similar protocol was used with the MGIA and BCG infection. Antibody (or PBS) was added to each tube in concentrations as indicated, in triplicates. The tubes were incubated for 96 hours at 37 ºC in a shaking incubator at 20 rpm. After incubation, tubes were centrifuged for 10 minutes at 2,000 g, and then 8ml sterile water was added per tube and the tubes were incubated for 10 minutes at room temperature. After blood lysis, tubes were spun at 2,000 g for 10 minutes, supernatant discarded and the pellet re-suspended in 1ml PBS. The samples were serially diluted and then plated onto OADC-supplemented 7H10 agar plates and incubated for 3 weeks at 37 ºC. For the depletion and/or blocking experiments in the MGIA, the experiment was performed as above but with the following modifications^24^. For the T cell depletion experiments, following a blood draw of 50 µl of the Human CD3 MicroBeads, CD4 Microbeads or CD8 MicroBeads (Miltenyi Biotec) were added separately into a 2ml diluted blood sample each and incubated for 30 minutes at 4 ºC. The LS columns (Miltenyi Biotec) were attached to the Midimacs and primed with 3ml PBS and 3ml RPMI-1640 sequentially. Next, 6ml of the diluted samples with the beads as above were added to the column. Control blood samples without CD3,CD4, or CD8 microbeads incubation were prepared under the same conditions as above. For the receptor blocking experiments, the Fc receptor antibodies anti-human CD32A (clone: 6C4; eBioscience) and anti-human CD16 (clone: 3G8; BioLegend) were used as the blocking antibodies for this assay. In the designated samples, 1 µg CD32 or 2 µg CD16 or both were used. For the MHC class II blocking experiments, 4 µg/ml mouse anti-human MHC class II antibody (clone Tu39, BD Biosciences catalogue # 555557) was used in the assay.

### Mouse infection assay

Monoclonal antibodies were injected via the intraperitoneal (IP) route into BALB/c mice (500 µg/mouse, unless otherwise indicated) 5 hours prior to aerosol infection by the Glas-col inhalation exposure system. PBS was injected as a control. Cultures of Mtb strain 165 were diluted to the concentration of 1×10^6^ CFU/ml (10 ml) for the aerosol infection. The mice were loaded into the basket of the inhalation exposure system with the lid secured, and bacterial suspension was loaded into the nebulizer. The set program of the inhalation exposure system was: preheating 15 minutes and nebulizing 30 minutes, cloud decay for 30 minutes, and decontamination for 15 minutes. 24 hours following infection, 3 of the PBS control mice were sacrificed and the lungs were removed and homogenized by MP Fastprep. The actual infection dose delivered to the lung was verified as between 100–200 CFU/mouse for all experiments. The other mice were sacrificed after 2 weeks, and lungs were homogenized and plated for the CFU load on OADC-supplemented 7H10 plates as above. The plates were incubated in a 37 °C incubator for around 3 weeks before colonies were counted.

### Complex preparation and crystallization

The Fabs of p4–36 or p4–170 were mixed with PstS1 at a molar ratio of 1:3 at 4 °C. The complex was purified by size exclusion chromatography with a Superdex 200 Increase 10/300 column running in 150 mM NaCl and 20 mM Tris-HCl pH 8.0. The peak fractions containing the complex were collected and concentrated for crystallization. The PstS1-Fab p4–36 crystals were grown at 18 °C by using the hanging-drop vapor diffusion method with 1 μl protein (6.2 mg/ml) mixed with 1 μl reservoir solution containing 0.2 M sodium iodide and 22% (w/v) polyethylene glycol 3350. The PstS1-Fab p4–170 was concentrated to ∼ 10 mg/ml, and the crystals were grown by using a similar method comprised of 0.2 M lithium sulfate, 0.1 M Tris pH 8.5, and 25% (w/v) polyethylene glycol 3350 at 18 °C. Crystals were soaked in a reservoir solution supplemented with 15% glycerol and flash frozen in liquid nitrogen for data collection.

### Data collection, structure determination and refinement

The diffraction data were collected on the BL17U beamline at the Shanghai Synchrotron Research Facility (SSRF). Indexing and integration were performed with the XDS software^39^, followed by scaling and merging with AIMLESS^40^. The structure was determined by molecular replacement using PHASER^41^. Manual building and adjustments of the structures were performed in COOT^42^. The structures were refined by using PHENIX^43^. Data collection and refinement statistics are listed in Table S2. Structural analyses of antibody-antigen contacts were assessed through CCP4i^44^ (Table S3 and S4). All structural representations were prepared through the use of the UCSF Chimera^45^.

### PstS1 mutation analysis

Site directed mutagenesis by PCR was used to introduce point mutations into recombinant PstS1 in the pMALp vector. The PCR products were cleaned with KIT PCR purification (Life Technologies), and the methylated parent DNA was digested using *dpnI* restriction enzyme (NEB). The restriction products were cleaned and transformed into DH5αF^−^ competent cells (Bio-Lab), after which the sequence was validated. The mutated PstS1 proteins were produced similarly to the procedure described above and tested for binding to p4-mAbs in ELISA.

## Extended Figures and Tables

**Extended Data Table 1: Patient clinical data**

**Extended Data Table 2: Data collection and refinement statistics**

**Extended Data Table 3: Fab p4–36/PstS1 interface**

**Extended Data Table 4: Fab p4–170/PstS1 interface**

**Extended Data Figure 1: Expression of Mtb antigens in E. coli fused to maltose binding protein. (a)** Schematic map of the pMALp-Mtb plasmid used as backbone for cloning of the selected Mtb antigens. Detailed descriptions of the cloning strategy and of the elements are shown on the right side of the figure. Gray – signal peptide; light pink – maltose binding protein (mbp); orange – TEV cleavage site; red – Mtb antigen, green – 6xHis; blue – Avi tag used for BirA biotinylation. **(b)** Protein SDS PAGE stain-free gel showing the different Mtb antigens fused to mbp after Ni affinity chromatography purification. The size of each corresponding protein is indicated.

**Extended Data Figure 2: V_H_ and V_L_ gene usage in PstS1-sorted memory B cells. (a)** Gene usage of the different V_H_ and V_L_ paired segments in the PstS-1 specific sorted cells. The color code is given in the lower part of the figure. **(b)** V_H_ gene frequency in single cells as oppose to clonal cells. Color code is the same as in (a).

**Extended Data Figure 3: Binding of anti-PstS1 mAbs to intact bacteria**. Reactivity to whole H37Ra-mCherry bacteria as determined by flow cytometry for four anti-mAbs, p4–163, p4–170, p4-36, and p4–141, and one isotype control mAb, MGO53.

**Extended Data Figure 4:Affinity maturation is necessary for the activity of p4–36 and p4–163 mAbs (a)** and **(b)** amino acid sequence alignment of the heavy chains and light chains of mAbs p4-163, p4–170, and p4–36 and their predicted germline versions, respectively. The alignment of Clone 1 antibodies (p4–163 and p4–170) is shown in the top panel, and the alignment of p4–36 in the bottom panel. The positions that are different from the germline are colored in light blue. **(c)** Binding by ELISA of the mature (mt) and the predicted germline (gl) versions of p4–36 (red) and p4–163 (blue), as well as their chimeras (mtHCglLC and glHCmtLC) to recombinant PstS1, Mtb-CDC1551 lysate **(d)**, and Mtb-H37Rv lysate **(e)**. MGO.53 serves as an isotype control.

**Extended Data Figure 5: Asymmetric unit of the PstS1-Fab p4–36 complex. (a)** Ribbon diagrams of two PstS1-Fab p4–36 heterodimers in the asymmetric unit of the crystal. The two heterodimers are colored cyan and yellow, respectively. **(b)** Structural superimposition of two PstS1-Fab p4–36 heterodimers in the asymmetric unit showing minor differences between the constant regions of the bound Fabs.

**Extended Data Figure 6: Comparisons of the epitopes recognized by p4–36 and p4–170. (a)** Ribbon (upper panel) and topology (lower panel) diagrams of PstS1 showing the epitope recognized by p4–36. The structural segments constituting the epitope are colored red. **(b)** Ribbon (upper panel) and topology (lower panel) diagrams of PstS1 showing the epitope recognized by p4–170. The structural segments constituting the epitope are colored red. Strands are represented as light green arrows and helixes are represented as blue cylinders in the topology diagrams.

**Extended Data Figure 7: Residues and CDRs of p4–36 and p4–170 involved in recognizing PstS1. (a)** Amino acid sequences of the p4–36 heavy chain variable region (VH) and light chain variable region (VL). The CDR regions in V_H_ and V_L_ are highlighted in purple and yellow, respectively. Residues in contacts with PstS1 are marked with red dots at the bottom of the sequence. **(b)** Ribbon diagrams of Fab p4–36 showing the CDRs and residues involved in binding PstS1. The CDRs of the heavy chain (cyan) and the light chain (hot pink) are colored purple and yellow, respectively. Positions of the residues involved in binding PstS1 are indicated by red balls. **(c)** Amino acid sequences of the p4–170 heavy chain variable region (VH) and light chain variable region (VL). The CDR regions in V_H_ and V_L_ are highlighted in purple and yellow, respectively. Residues in contacts with PstS1 are marked with red dots at the bottom of the sequence. **(d)** Ribbon diagrams of Fab p4–170 showing the CDRs and residues involved in binding PstS1. The CDRs of the heavy chain (cyan) and the light chain (hot pink) are colored purple and yellow, respectively. Positions of the residues involved in binding PstS1 are indicated by red balls.

## Data Availability

all data associated with a paper is available, and can be accessed

## Acknowledgments

This study was funded by Israeli Science Foundation grant 1422/18 to N. T. Freund and Israeli Innovation Authority grant 65029 to N. T. Freund, as well as a National Natural Science Foundation of China grant (81661128044) to B. Javid, who is a Welcome Trust Investigator. We thank all members of the Freund Laboratory at Tel Aviv University for fruitful discussions and technical help. We thank N. Ben-Shalom for help with the flow cytometry analysis and figures. We thank O. Freund for his ideas, his support, and his assistance with computational antibody sequence analysis. We thank T. Pliss and A. Tarn for their support throughout the project. We thank M. C. Nussenzweig and J. Ernst for critical reading and for their valuable feedback.

## Authors contributions

A. Watson, H. Li and B. Ma designed and carried out the experiments, and analyzed the data. A. Watson and R. Weiss conducted the single-cell sorting, antibody sequencing and cloning, ELISA, and Flow Cytometry assays. H. Li preformed the whole blood assays and the in vivo assays. Z. Wang and F. Du. assisted H. Li with the pathogenic Mtb experiments. B. Ma preformed all the structural analyses and data collections and prepared the structural figures. D. Bendayan recruited the ATB patients. E. Pinco was responsible for sample collection and handling. L. Abramovich carried out the THP-1 assays. M. Bar-Oz and D. Barkan grew H37Ra bacteria and helped with the infections of THP-1 cells. M. Mordekovich expressed the anti-PstS1 mAbs. J. Rybniker conducted confocal microscopy and advised throughout the project. H. Huang and Y. Lu provided biosafety facilities and helped supervise pathogenic Mtb experiments. N. T. Freund, B. Javid, and Y. Xiang designed the experiments, analyzed the data, prepared the figures, and wrote the manuscript with input from the other authors.

